# Linking Free Text Documentation of Functioning and Disability to the ICF with Natural Language Processing

**DOI:** 10.1101/2021.08.19.21262308

**Authors:** Denis Newman-Griffis, Jonathan Camacho Maldonado, Pei-Shu Ho, Maryanne Sacco, Rafael Jimenez Silva, Julia Porcino, Leighton Chan

## Abstract

**Background:** Invaluable information on patient functioning and the complex interactions that define it is recorded in free text portions of the Electronic Health Record (EHR). Leveraging this information to improve clinical decision-making and conduct research requires natural language processing (NLP) technologies to identify and organize the information recorded in clinical documentation.

**Methods:** We used NLP methods to analyze information about patient functioning recorded in two collections of clinical documents pertaining to claims for federal disability benefits from the U.S. Social Security Administration (SSA). We grounded our analysis in the International Classification of Functioning, Disability and Health (ICF), and used the ICF’s Activities and Participation domain to classify information about functioning in three key areas: Mobility, Self-Care, and Domestic Life. After annotating functional status information in our datasets through expert clinical review, we trained machine learning-based NLP models to automatically assign ICF categories to mentions of functional activity.

**Results:** We found that rich and diverse information on patient functioning was documented in the free text records. Annotation of 289 documents for Mobility information yielded 2,455 mentions of Mobility activities and 3,176 specific actions corresponding to 13 ICF-based categories. Annotation of 329 documents for Self-Care and Domestic Life information yielded 3,990 activity mentions and 4,665 specific actions corresponding to 16 ICF-based categories. NLP systems for automated ICF coding achieved over 80% macro-averaged F-measure on both datasets, indicating strong performance across all ICF categories used.

**Conclusions:** NLP can help to navigate the tradeoff between flexible and expressive clinical documentation of functioning and standardizable data for comparability and learning. The ICF has practical limitations for classifying functional status information in clinical documentation, but presents a valuable framework for organizing the information recorded in health records about patient functioning. This study advances the development of robust, ICF-based NLP technologies to analyze information on patient functioning, and has significant implications for NLP-powered analysis of functional status information in disability benefits management, clinical care, and research.

## 1. Introduction

Understanding a person’s functioning requires a multifaceted picture of the complex interactions between the person and the world around them. The International Classification of Functioning, Disability and Health (ICF) (1) conceptualizes these interactions as between a person’s health condition(s), body structures and functions, activities and participation, and both environmental and personal contextual factors. In order to fully capture the multifactorial nature of functional outcomes and a person’s experience of their functioning, providers primarily turn to free text documentation in the Electronic Health Record (EHR) (2–4). While the flexibility of free text presents a barrier to standardization in the EHR, limiting comparability across patients and opportunities for data-driven learning in modern health systems (5), the expressivity of natural language is key to capturing the nuances of functioning as it is experienced in the patient’s life (6). For example, two patients reporting moderate limitations in walking may experience them in entirely different ways: one may describe arthritic stiffness in their knees that causes manageable discomfort in navigating employment in an office, while another patient’s chronic low back pain makes their hiking hobby no longer viable. These differences in experience, which inform both therapeutic interventions and the patient’s perception of their own functioning, are difficult to capture in standardized instruments but can be easily described in natural language.

How to navigate the tradeoff between flexibility in clinical documentation and standardization for comparability and learning? We explore the use of natural language processing (NLP) systems, grounded in the ICF, to index and organize information about functioning and disability in free text clinical records, enabling a measure of standardization without sacrificing the details of patient experience. NLP can be used to identify, organize, and retrieve information from free text documents for use in clinical decision making and research (7,8). NLP shows growing promise for capturing and analyzing information on functioning: Kukafka et al. (9) developed an early system for coding rehabilitation discharge summaries to identify activities including eating, dressing, and toileting, and NLP has since been used for a variety of purposes including locating functional status documentation in oncology notes (10), identifying potential wheelchair use (11), and detecting functional outcomes of geriatric syndrome (12). We have previously developed NLP methods to identify activity mentions describing mobility functioning in clinical notes (13–15), and to link these activity mentions to the Mobility chapter of the ICF’s Activities and Participation domain (16).

This study investigated NLP methods for automatically coding documentation of key domains of functioning to the ICF, and evaluated their performance on coding medical records associated with claims for federal disability benefits submitted to the U.S. Social Security Administration (SSA). We adapted our previous work on Mobility information to expand to information from the Self-Care and Domestic Life chapters of the ICF’s Activities and Participation domain. Together with Mobility, these domains align with the majority of Activities of Daily Living (ADLs)—fundamental activities frequently considered in therapeutic patient assessment, such as dressing, hygiene, eating, and ambulation—(17,18), and account for 11 of the 18 items in the Functional Independence Measure (FIM)—a tool for assessing a patient’s degree of independence, commonly used in assessing rehabilitation outcomes (19). Thus, NLP methods to automatically identify activities in these three ICF chapters have significant potential for use in clinical information systems.

The remainder of this article is organized as follows. In the Materials and Methods section, we describe the medical records we analyzed from SSA disability benefits claims and present the NLP methods used for linking information about patient function in these records to relevant categories in the ICF. The Results section presents our experimental findings and an analysis of successes and challenges in coding clinical data with the ICF. The Discussion section outlines implications from our work, including challenges for applying the ICF in coding clinical notes, opportunities for NLP impact in the SSA disability adjudication process and in broader clinical information systems, and limitations of the study.

## 2. Materials and Methods

Our study involved the development and evaluation of machine learning-based statistical models for linking descriptions of Mobility, Self-Care, and Domestic Life functioning in free text clinical documentation to relevant categories in the ICF. While we consider automated assignment of the qualifier component of ICF codes out of scope for this study, and use two-level classification categories for the output of our NLP systems, we refer to this process as *ICF coding* to align it with prior literature on automated medical coding systems. We use the term *functional status information* (FSI) to refer to information about patient functioning, including specific observations in activity mentions).

### 2.1. Data sources and use of the ICF

Our primary data source for this study was free text medical records collected by SSA in the process of adjudicating federal disability benefits claims. During the adjudication process of an individual’s claim, SSA may obtain records from that individual’s prior medical encounters in order to collect medical evidence related to the disability claim. These records are reviewed by expert adjudicators at SSA to identify appropriate evidence to support the claim decision, such as impairment history and severity, relationship to work requirements, etc. The volume of these records is substantial, with each claim having potentially hundreds or thousands of pages of associated medical records, presenting a significant opportunity for NLP methods to assist in evidence review by automatically identifying relevant information.

We used two types of medical documents in the study. (1) Consultative Examination (CE) reports are written by a medical expert commissioned by SSA to examine a claimant in-depth as part of the claim adjudication process. (2) EHR data are provided directly to SSA by health providers pursuant to a disability benefits claim. Both types of documents are frequently submitted to SSA as faxed or scanned documents, and thus require Optical Character Recognition (OCR) to convert them to text for NLP analysis. All documents used in this study were converted to text using the Nuance OmniPage™ (now Kofax OmniPage Ultimate™) OCR software.

We selected the ICF, and the Activities and Participation domain in particular, as our framework for identifying functioning information in these documents. We chose the ICF due to its role as an internationally recognized coding system for functioning, and our familiarity with it (6,15,16). SSA assesses function as part of the claim adjudication process, including assessment of residual functional capacity for individuals applying for disability benefits, examining both physical and mental function. We identified the Mobility, Self-Care, and Domestic Life chapters of the ICF as being most relevant to this process and the types of functioning documented most frequently in the data we reviewed. As noted in the Introduction, these chapters are also closely aligned with commonly used ADL measures and the FIM, making them particularly relevant types of information to study for a broad range of information needs in rehabilitation. We use title case in this article to refer to Mobility, Self-Care, and Domestic Life information, as defined by the ICF, to distinguish from the more general uses of these terms.

#### 2.1.1. Document collections for annotation

We identified two sets of medical documents from SSA to annotate for Mobility, Self-Care, and Domestic Life FSI. Both datasets for annotation were drawn from adult disability benefits claims with a decision issued in 2016-2018, primarily related to musculoskeletal, neurological, or mental impairments.

Following our prior work on analyzing Mobility information (15), we identified 300 CEs likely to contain descriptions of Mobility functioning. We ensured that each CE corresponded to a different claimant in order to control for cross-document correlation from an individual claimant.

An additional 350 documents were then selected to annotate for Self-Care and Domestic Life information. The documents were selected from the same overall set of claims as the Mobility documents, but we ensured that the specific claims used in annotation were disjoint between the two datasets. As the concepts of Self-Care and Domestic Life are highly intertwined and often discussed together in clinical notes—e.g., eating (Self-Care) and preparing meals and cleaning (Domestic Life)—we chose to annotate for these chapters jointly (referred to in the remainder of the article as “Self-Care/Domestic Life”). Annotated documents included both CEs and EHR data; no two documents of the same type were included for any individual claimant.

#### 2.1.2. SSA document collection for computational language modeling

A further set of 65,514 documents collected by SSA were used for machine learning of statistical models of clinical language as used in the SSA setting (as detailed in the “Text representation with word embeddings” section below). Many documents included in this collection included notes from multiple clinical encounters during a patient’s history with a particular healthcare provider. Each “document” was thus much longer on average than a single clinical note, with a median document length of 3,476 words. These documents were sampled by SSA separately from the documents used for annotation, using a broader set of criteria to enhance diversity of the data: adult claims adjudicated based on musculoskeletal, neurological, or mental impairments, with a decision issued during 2013-2018, drawn from multiple states around the U.S. We confirmed that no documents selected for Mobility or Self-Care/Domestic Life annotations were included in this collection.

### 2.2. Annotation process

Annotation of SSA documents for FSI regarding Mobility and Self-Care/Domestic Life was performed in a multi-stage process, illustrated in Figure 1. Mobility information was annotated using guidelines developed in previous work (15); we adapted this existing process to develop new guidelines for Self-Care/Domestic Life information. We developed the annotation guidelines via an iterative process among the annotators (JCM, PSH, MS, RJS), involving team annotation and discussion to refine a schema for representing Self-Care/Domestic Life information and develop clear guidelines for how to annotate for it in free text. After guideline development, the annotators jointly annotated a small set of documents (50 for the new Self-Care/Domestic Life guidelines, and 16 to further validate the existing Mobility guidelines in SSA data), and Inter-Annotator Agreement (IAA) was calculated (IAA values are reported with other dataset statistics in the Results section).

**Figure 1.**
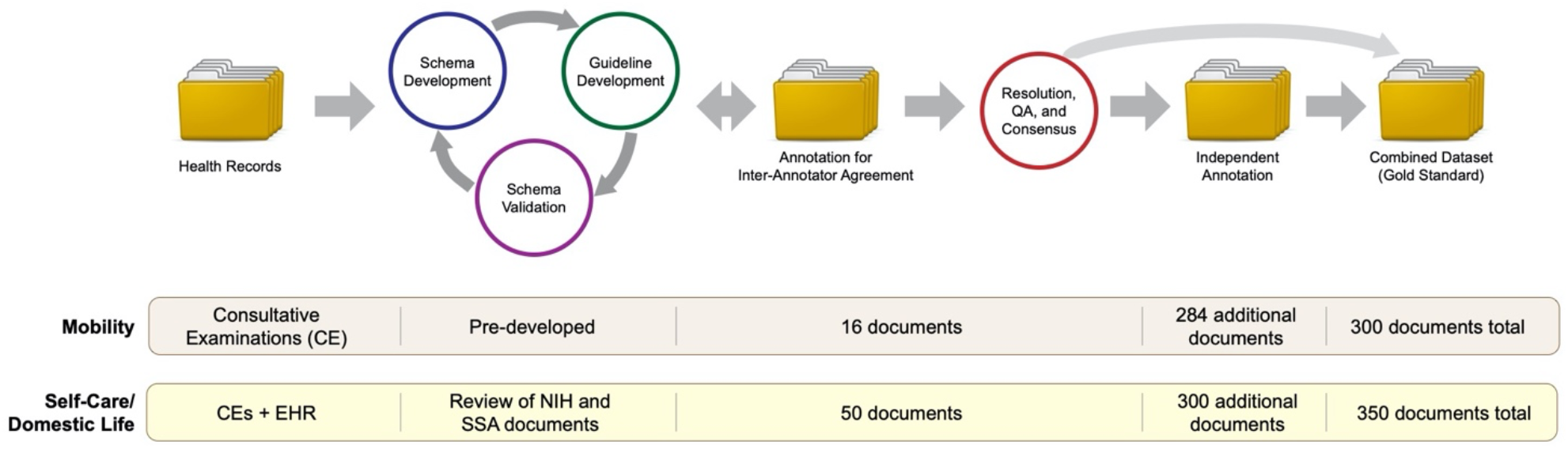
Flowchart illustration of annotation process. Data sources and document counts are provided for Mobility and Self-Care/Domestic Life annotations separately.

Following standard practice in annotating for text spans (20,21), we calculated IAA using the F-1 measure. Disagreements were then resolved by joint meetings among the annotators to produce a final consensus version of the jointly-annotated documents. Finally, each individual annotator annotated a further set of documents independently, which were then combined with the consensus annotations to produce the final “gold standard” annotated corpus.

When annotating a document, the first step in our process was to identify *activity mentions*, which we operationalized as self-contained spans of text describing a person’s functioning within the scope of the relevant ICF Activities and Participation chapters. Within each activity mention, we then identified each distinct *action* referred to, operationalized as a distinct activity defined by one of the ICF categories within the relevant chapters of the two-level ICF classification (or an activity of similar granularity not specifically captured in the ICF, e.g., “do household chores”). These categories are represented using the ICF format of the letter *d* (indicating the Activities and Participation domain) followed by three digits: a one-digit chapter identifier and a two-digit category identifier (e.g., d450 indicates the *Walking* category in Chapter 4 *Mobility*). We refer to these as *second-level categories* to distinguish them from the more specific subcategories in the detailed classification (e.g., d4501 *Walking long distances*).

Each of the identified action components (which we denote with a capitalized *Action* for the remainder of this article, for clarity) within an activity mention was then assigned the second-level ICF category best representing the activity described. We excluded the “other specified” and “unspecified” ICF categories, such as d598 *Self-care, other specified* and d599 *Self-care, other unspecified*, from use in annotation due to their ambiguity. In cases where an Action component referred to an activity for which no specific ICF category was appropriate (e.g., “doing household tasks”), or when multiple categories could apply (e.g., “denies difficulty with ADLs”), a label of “Other” was used. Figure 2 provides an illustrated example of Self-Care/Domestic Life activity mentions, including one with two Action components.

**Figure 2.**
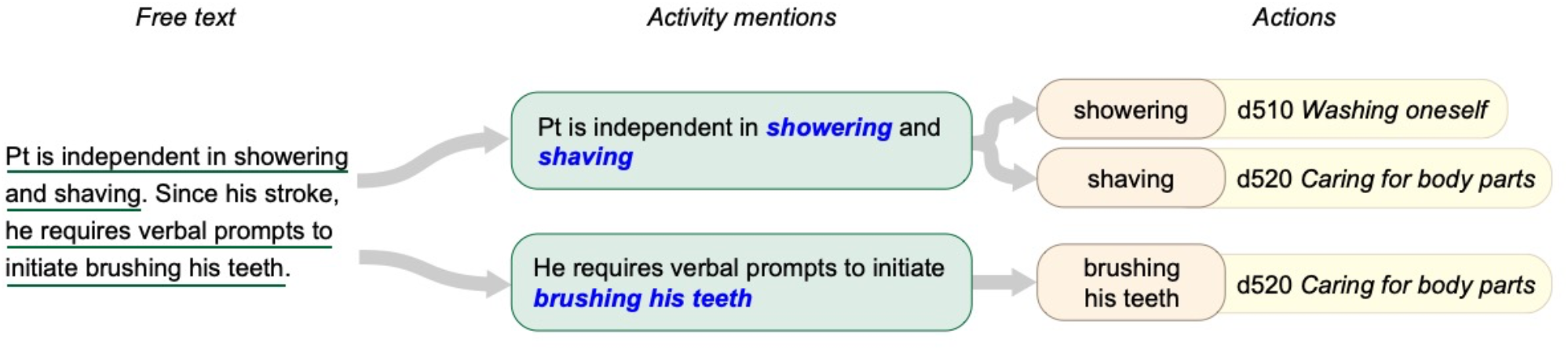
Structure of annotations for functional status information. Free text is annotated to identify activity mentions describing specific observations. Each activity mention may include one or more Action components, which can be mapped to second-level ICF categories.

The focus of annotation was on observations or descriptions of specific, volitional activities performed by the patient within the specific domains of interest. We therefore excluded the following types of information about functioning: (1) hypothetical statements (e.g., “her sleep is better if she takes medication); (2) education given by the provider (e.g., “Patient educated on how he can attempt to dress his lower body in bed”); and (3) references to habitual activity in the context of work duties (e.g., “his job at the hotel involves doing laundry and cleaning guest rooms”).

#### 2.2.1. Patient engagement in medication management and non-pharmacological therapies as categories of self-care

The documents reviewed for Self-Care/Domestic Life guideline development included frequent discussions of patients’ active engagement in the therapeutic process, including adherence to medication management regimens and participation in non-pharmacological therapies. While these mentions provided valuable evidence of distinct kinds of patient engagement in self-care, they were not reflected by ICF categories more specific than d570 *Looking after one’s health*. To more accurately capture—and differentiate between—these frequent topics, we added two additional Action labels based on codes in the Systematized Nomenclature of Medicine Clinical Terms set (SNOMED CT). We used *Manage medication* (SNOMED CT code 285033005) to refer to anything related to compliance with medications such as the ability to store medications, obtain medications, taking the medications, etc. This label also included the mismanagement of medication (e.g., forgetting to take prescribed medications). We used *Therapy* (SNOMED CT code 709007004) to refer to attending or otherwise engaging in non-pharmacological therapies such as addiction treatment programs, physical therapy, occupational therapy, cognitive behavioral therapy, psychological therapy, and anger management. We did not use these labels to annotate the therapeutic interventions themselves, which are out of scope of the ICF. Thus, while a mention of a patient attending physical therapy was annotated as a *Therapy* activity mention, a mention of a physical therapy appointment with no indication of whether the patient attended or not did not provide evidence of self-care, and was not annotated.

### 2.3. Methods for automated ICF coding

We experimented with two strategies to develop computer methods to automatically assign ICF categories to Mobility and Self-Care/Domestic Life activity mentions. In our prior work (16), we explored a variety of methods for ICF coding, including both *classification*—identifying the group of samples a given activity mention is most similar to—and *candidate selection*—identifying which ICF category a given activity mention is most similar to—approaches, for Mobility information only. In this study, we evaluated the best-performing classification and candidate selection models from this prior work on the SSA datasets we developed for Mobility and Self-Care/Domestic Life. Our overall process is illustrated in Figure 3.

**Figure 3.**
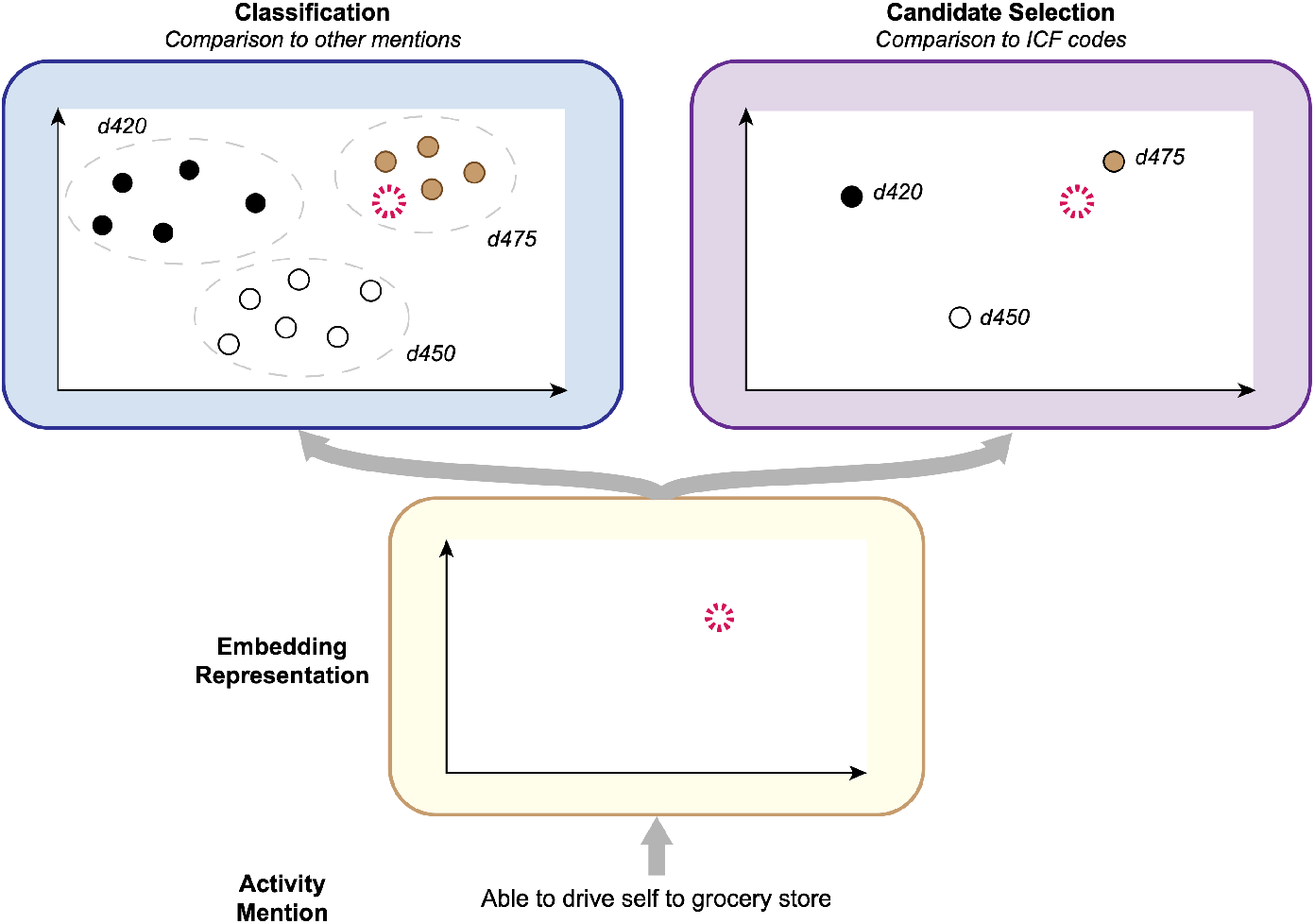
Conceptual illustration of ICF coding process. Given an activity mention, an embedding representation of the report is calculated, and then compared to (a) other activity mentions, in Classification; or (b) available ICF categories, in Candidate Selection.

#### 2.3.1. Text representation with word embeddings

Given an activity mention, we calculated a numeric representation of the text using word embedding features. In word embedding models, each word and phrase is represented mathematically using a vector of *n* real numbers—frequent values for *n* include 100, 300, and 768—with the property that words that are similar in meaning generally have similar numeric representations (22). These models are fundamental resources for modern NLP methods. Our prior work demonstrated that word embedding features alone were more informative for ICF coding than features indicating the presence and/or frequency of specific words (referred to as *lexical features*) or combined embedding and lexical features (16); we therefore used word embedding features alone in this study. We experimented with two methods for word embedding:

- In *static* embeddings, each unique word is represented by a single vector. Thus, for example, every occurrence of the word *patient* is represented within the model using the same set of real numbers. We used FastText (23), a commonly used method that integrates sub-word information into embedding learning to better capture morphological patterns.
- In *contextualized* embeddings, each word is represented by a single vector conditioned on the context it appears in; thus, the word “cold” in “patient described <u>cold</u> symptoms” and “applied a <u>cold</u> pack” is represented using different vectors of real numbers for each case. This provides additional context-sensitivity in how the model represents text content. We used BERT (24), a recent embedding model that has rapidly become the *de facto* standard for text representation in NLP.

The parameters of both static and contextualized embedding models (i.e., the values used to represent words and phrases) are typically estimated prior to their usage in any specific NLP task (e.g., our ICF coding application), based on a large sample of natural language (referred to as a *corpus*). Different corpora may be chosen for different purposes—e.g., estimating an embedding model using the text of PubMed abstracts provides useful representations for analyzing scientific literature, while using the text of clinical notes provides more useful representations for clinical applications. We therefore experimented with multiple corpora to estimate our word embedding models (referred to in machine learning as *model training*), each of which reflects different tradeoffs between corpus size and representativeness for the target task. These corpora are summarized in Table 1.

**Table 1.** Free text corpora used to train word embedding models for text representation. MIMIC-III was used to train both FastText and BERT models; NIHCC and SSA were used for FastText embeddings only.

| Training corpus | Number of notes | Number of words (approx.) | Data description |
| --- | --- | --- | --- |
| MIMIC | 2,083,180 | 497 million | Critical care admissions (26). Most commonly used corpus for language modeling in clinical NLP. |
| NIHCC | 63,605 | 11.8 million | Physical therapy and occupational therapy encounters, used in our prior work on coding Mobility information to the ICF (16). |
| SSA | 65,514 | 664 million | Clinical data associated with disability benefits claims submitted to SSA. New in this study. |

For static word embeddings, we experimented with three clinical corpora for training embedding models. In each case, document texts were broken down into individual words (tokenized) with the spaCy software (25), and the following processing steps were applied to normalize out aspects of the text irrelevant to our language modeling goal: all words were converted to lowercase, all numbers were normalized to “[NUMBER]”, all URLs were normalized to “[URL]”, and all dates and times were normalized to “[DATE]” and “[TIME]” respectively. The FastText software (version 0.2.0) was used with the skipgram algorithm, 300-dimensional embeddings, and all other settings at default to train embeddings on the following three corpora:

- **MIMIC**: Approximately 2 million free text notes included in the publicly-available Medical Information Mart for Intensive Care (MIMIC) critical care database, version 3 (26). Notes are associated with admissions to ICU units of Beth Israel Deaconess Medical Center in Boston between 2001-2012, and are commonly used for language modeling in clinical NLP research.
- **NIHCC**: Over 63,000 free text notes from 10 years of Physical Therapy and Occupational Therapy encounters in the Rehabilitation Medicine Department of the NIH Clinical Center, collected and used for calculating word embedding features in our previous work (16).
- **SSA**: Over 65,000 free text notes associated with disability claims processed by SSA within a five-year period (as described in the “SSA document collection for language modeling” section above).

Contextualized embedding models require significant computing power to train on new data, and pre-trained models are typically used to generate text features. We used the clinicalBERT model released by Alsentzer et al. (27), which was trained on MIMIC clinical notes, and produces 768-dimensional word embeddings.

##### 2.3.1.1. Action oracle

As illustrated in Figure 2, activity mentions are complex statements including multiple pieces of information. Thieu et al (15) define sub-components of activity mentions, including (1) a source of Assistance—typically a device, person, or structure in the physical environment used in activity performance; (2) a Quantification—an objective measure of functional performance, such as distance or time; and (3) one or more specific Actions being performed, which correspond to defined activities in the ICF Activities and Participation domain. For example, the activity mention “Pt ambulated 300’ in clinic with rolling walker” includes the Action component “ambulated”, the Assistance component “with rolling walker”, and the Quantification component “300’”. Action components are annotated with the second-level ICF categories which the NLP systems described in this study are designed to assign.

Prior work on extracting activity mentions from free text (13,14) did not include extraction of the Action sub-components. However, as NLP methods for functional status information continue to develop, more complex models that reflect the semantic structure of activity mentions will be needed. We therefore evaluated the ICF coding models in this study in two settings: (1) an *Action oracle* setting, in which both an activity mention and the location of an Action component within it (i.e., where in the activity mention’s text span the Action is found) are input to the ICF coding model; and (2) a non-oracle setting in which only the activity mention is provided (reflecting the technologies so far developed for extracting activity mentions).

#### 2.3.2. Classification

In classification approaches, a mathematical representation is calculated for each activity mention using word embedding features, and a predictive model is trained to assign an ICF category to each Action component based on its similarity to previously-observed samples labeled with each ICF category. We adopted the best-performing classification model from our prior work (16), a Support Vector Machine (28) using word embedding features as input. Given an input activity mention, we calculated its embedding features in one of four ways:

- **Static embeddings, no Action oracle**: the activity mention is represented by averaging the word embeddings of each word in the mention.
- **Static embeddings, with Action oracle**: two averaged embeddings are calculated: (1) the averaged embedding for the words in the Action component; and (2) the average of other all words in the activity mention. These are concatenated, i.e., combined into a single, longer vector, to produce the final representation.
- **Contextualized embeddings, no Action oracle**: the activity mention is represented as the averaged context-sensitive embeddings for each of its words.
- **Contextualized embeddings, with Action oracle**: as the contextualized embeddings of words in the Action component already reflect information about the full activity mention, we averaged the embeddings of Action component words only.

#### 2.3.3. Candidate selection

In the candidate selection approach, an embedding representation is calculated for each activity mention, and is then compared to embedding representations of each of the available ICF categories to identify which category the given mention is most similar to. We adopted the best-performing candidate selection model from our prior work (16), consisting of a Deep Neural Network (DNN) that operates as follows:

1. The model takes as input an activity mention embedding and embedding representations of the ICF categories that could be assigned to it (i.e., all Mobility categories or all Self-Care/Domestic Life categories).
2. These embeddings are all fed into a DNN to calculate new embedding representations of the candidate ICF categories, conditioned on this specific activity mention.
3. The conditional ICF category embeddings are compared to the activity mention embedding using the cosine similarity measure, and the category with highest similarity is chosen as the model output.

Embedding features of activity mentions were calculated using the strategies described in the “Classification” section. Embedding representations of ICF categories were calculated as the averaged embeddings of each word in the definition of the category presented in the ICF, using both static and contextualized embeddings. For the “Other” label, the following definitions were used: “Mobility other or unspecified” for Mobility, and “Self-care or domestic life other or unspecified” for Self-Care/Domestic Life. For the added *Therapy* and *Manage medication* labels, we used the names of the corresponding SNOMED CT codes (“Ability to manage medication” and “Compliance behavior to therapeutic regimen”, respectively). Further details of the model are presented in (16). Following our prior work, we used a 3-layer DNN with hidden layer size 300 when using static embedding features without the Action oracle, a 3-layer DNN with layer size 600 when using static embeddings with the Action oracle (to match the dimensionality of the concatenated activity mention and Action component embeddings), and a 1-layer DNN with layer size 768 when using BERT embedding features (for which vector dimensionality does not change with the Action oracle).

### 2.4. Experimental procedure

Prior to machine learning experiments, each dataset was split at the document level into training data, for training the machine learning models, and test data for evaluating them. Test documents were sampled to include at least 20% of the samples for each ICF category. Statistical significance testing was performed using the bootstrap resampling method with 1000 replicates, which is commonly used to analyze performance metrics in NLP research (29,30).

#### 2.4.1. Development experiments

Training data was further split into ten folds for development experiments to select the best word embedding method for classification and candidate selection approaches. For development experiments, cross validation was used: models were trained on nine folds (90% of the training data) and evaluated on the held-out tenth fold, and this process was then repeated to evaluate on each of the ten folds, with model performance being averaged across the folds to calculate final values. Model performance was calculated using F-1 score (20), calculated as the harmonic mean between precision (positive predictive value) and recall (sensitivity). F-1 score was calculated for each ICF category in each dataset, and averaged across categories to calculate macro F-1. The embeddings producing highest macro F-1 on the development experiments were chosen to use for the main experiments.

#### 2.4.2. Main experiments and model evaluation

Once final word embeddings were chosen, an additional classification and candidate selection model was trained for each of the Mobility and Self-Care/Domestic Life datasets, using all of the training data. These models were then evaluated on the held-out test documents, with performance measured using F-1 for each individual ICF category, and overall performance calculated as macro-averaged F-1 score.

## 3. Results

### 3.1. Annotated datasets

Table 2 presents overall statistics of the two SSA datasets annotated for functional status information. Several of the documents selected for annotation were omitted after conversion to text with the OCR software due to failures in the OCR conversion, resulting in a total of 289 documents annotated for Mobility, and 329 documents annotated for Self-Care/Domestic Life. The majority of documents were found to contain descriptions of the target types of functioning: 251/289 (87%) of Mobility documents and 285/329 (87%) of Self-Care/Domestic Life documents contained at least one activity mention pertaining to the relevant ICF chapters. Each activity mention could contain zero, one, or more than one Action component; a total of 3,176 Actions were annotated for Mobility and 4,665 for Self-Care/Domestic Life. Only 132 Mobility activity mentions (5.4% of the total) and 134 Self-Care/Domestic Life activity mentions (3.4% of the total) were found to not contain any specific Action components. Inter-annotator agreement (IAA) was found to be 0.778 F-1 for Mobility and 0.695 F-1 for Self-Care/Domestic Life, comparable to IAA calculated in our previous study on annotating Mobility information in clinical reports (15). ICF coding has previously been found to be high agreement for resources and goals as well as specific problems (31). The two datasets are described in greater detail in the following sections.

**Table 2.**
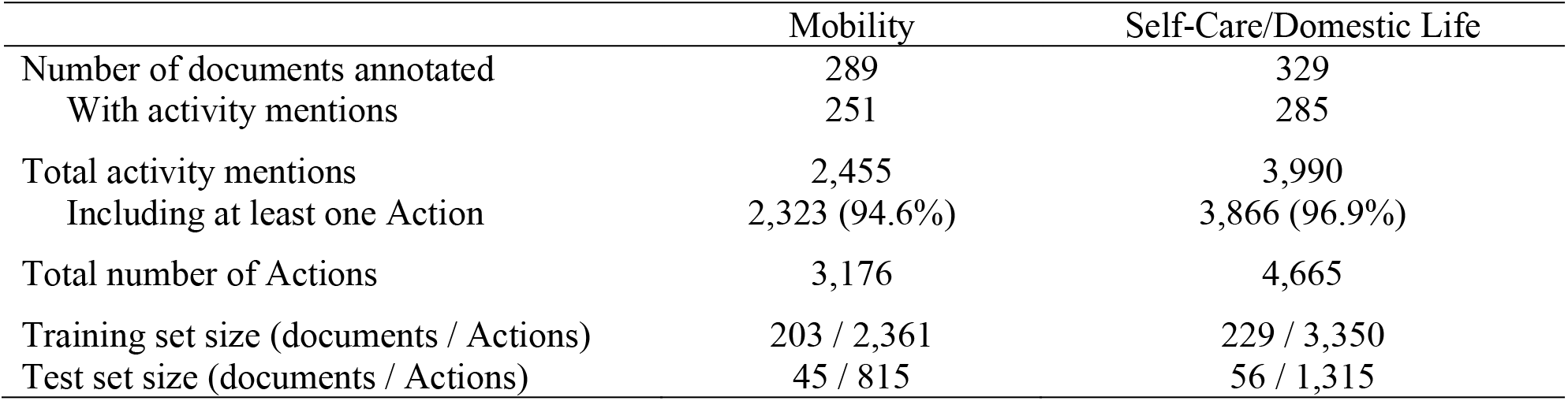
Datasets of documents annotated for functional status information, drawn from U.S. Social Security Administration disability benefits cases. Separate sets of documents were annotated for Mobility (ICF Activities and Participation Chapter 4) and Self-Care/Domestic Life (ICF Activities and Participation Chapters 5 and 6).

|  | Mobility | Self-Care/Domestic Life |
| --- | --- | --- |
| Number of documents annotated | 289 | 329 |
| With activity mentions | 251 | 285 |
| Total activity mentions | 2,455 | 3,990 |
| Including at least one Action | 2,323 (94.6%) | 3,866 (96.9%) |
| Total number of Actions | 3,176 | 4,665 |
| Training set size (documents / Actions) | 203 / 2,361 | 229 / 3,350 |
| Test set size (documents / Actions) | 45 / 815 | 56 / 1,315 |

#### 3.1.1. Mobility dataset

A total of twelve unique second-level ICF categories were used for annotating Mobility information; Table 3 lists the frequency of each of these categories in the annotated dataset, together with the “Other” category. Of the categories in the Mobility chapter, only d480 *Riding animals for transportation* was not observed in the annotation process. d465 *Moving around using equipment* was excluded from annotation, as the use of equipment was annotated using Assistance components of Mobility activity mentions; d455 *Moving around* was used instead. The most frequent categories were d450 *Walking* (23.0% of Actions), d410 *Changing basic body position* (17.6% of Actions), and d415 *Maintaining a body position* (16.0% of Actions). Only d420 *Transferring oneself*, d435 *Moving objects with lower extremities*, and d460 *Moving around in different locations* were observed fewer than 100 times. A total of 123 samples (3.9% of Actions) were found that could not be mapped to a single appropriate second-level ICF category. These included Actions which could map to multiple categories, such as “The patient is able to <u>ambulate</u> in the hallway and stairs” (which can refer to both d450 *Walking* and d460 *Moving around in different locations*), and Actions which were too vague to map to any specific categories, such as “The patient cannot <u>manage/negotiate stairs</u>”.

**Table 3.** ICF category descriptions and frequencies for Mobility dataset (3,176 samples total). Categories are ordered by frequency in the dataset. Sample count and relative distribution between training data (203 documents, 2361 samples) and test data (45 documents, 815 samples) are given for each category. Descriptions given are the preferred name of each category in the ICF.

| Mobility Category | Description | Frequency | % of All Samples | Training Samples | Test Samples |
| --- | --- | --- | --- | --- | --- |
| d450 | Walking | 730 | 23.0% | 559 (77%) | 171 (23%) |
| d410 | Changing basic body position | 560 | 17.6% | 419 (75%) | 141 (25%) |
| d415 | Maintaining a body position | 508 | 16.0% | 385 (76%) | 123 (24%) |
| d440 | Fine hand use | 319 | 10.0% | 247 (77%) | 72 (23%) |
| d430 | Lifting and carrying objects | 244 | 7.7% | 167 (68%) | 77 (32%) |
| d475 | Driving | 215 | 6.8% | 165 (77%) | 50 (23%) |
| d445 | Hand and arm use | 163 | 5.1% | 104 (64%) | 59 (36%) |
| d455 | Moving around | 147 | 4.6% | 99 (67%) | 48 (33%) |
| Other | Mobility-related activities for which no specific ICF category could be identified | 123 | 3.9% | 96 (78%) | 27 (22%) |
| d470 | Using transportation | 103 | 3.2% | 80 (78%) | 23 (22%) |
| d460 | Moving around in different locations | 55 | 1.7% | 34 (62%) | 21 (38%) |
| d435 | Moving objects with lower extremities | 5 | 0.2% | 4 (80%) | 1 (20%) |
| d420 | Transferring oneself | 4 | 0.2% | 2 (50%) | 2 (50%) |

#### 3.1.2. Self-Care/Domestic Life dataset

Thirteen distinct second-level ICF categories (seven from Chapter 5 *Self-Care*, six from Chapter 6 *Domestic Life*) were used in data annotation, together with the added labels of *Manage medication* and *Therapy* and the “Other” category. Table 4 lists the observed frequency of each of these labels in the dataset. The most frequent category was d570 *Looking after one’s health*, accounting for 43.6% of the samples by itself. Five categories (d530 *Toileting*, d560 *Drinking*, d610 *Acquiring a place to live*, d650 *Caring for household objects*, and d660 *Assisting others*) occurred fewer than 100 times. A total of 175 samples were found that could not be mapped to a single appropriate second-level ICF category, such as “The patient is independent with <u>ADLs</u>” (which includes multiple Self-Care activities).

**Table 4.** ICF category descriptions and frequencies for Self-Care/Domestic Life dataset (4,665 samples total). Categories are ordered by frequency in the dataset. Sample count and relative distribution between training data (229 documents, 3350 samples) and test data (56 documents, 1315 samples) are given for each category. Descriptions given are the preferred name of each category in the ICF.

| Self-care/<br>Domestic Life<br>Category | Description | Frequency | % of All Samples | Training Samples | Test Samples |
| --- | --- | --- | --- | --- | --- |
| d570 | Looking after one's health | 2,032 | 43.6% | 1496 (74%) | 536 (26%) |
| Manage medication | Ability to manage medication (SNOMED CT code 285033005) | 520 | 11.1% | 359 (69%) | 161 (31%) |
| d540 | Dressing | 353 | 7.6% | 268 (76%) | 85 (24%) |
| d520 | Caring for body parts | 312 | 6.7% | 228 (73%) | 84 (27%) |
| d640 | Doing housework | 297 | 6.4% | 205 (69%) | 92 (31%) |
| d630 | Preparing meals | 222 | 4.8% | 165 (74%) | 57 (26%) |
| Other | Self-Care/Domestic Life activities for which no specific ICF category could be identified | 174 | 3.7% | 127 (73%) | 47 (27%) |
| Therapy | Compliance behavior to therapeutic regimen (SNOMED CT code 709007004) | 143 | 3.1% | 103 (72%) | 40 (28%) |
| d620 | Acquisition of goods and services | 142 | 3.0% | 101 (71%) | 41 (29%) |
| d510 | Washing oneself | 121 | 2.6% | 90 (74%) | 31 (26%) |
| d550 | Eating | 102 | 2.2% | 57 (56%) | 45 (44%) |
| d560 | Drinking | 82 | 1.8% | 60 (73%) | 22 (27%) |
| d660 | Assisting others | 79 | 1.7% | 46 (58%) | 33 (42%) |
| d650 | Caring for household objects | 40 | 0.8% | 24 (60%) | 16 (40%) |
| d530 | Toileting | 29 | 0.6% | 15 (52%) | 14 (48%) |
| d610 | Acquiring a place to live | 17 | 0.3% | 6 (35%) | 11 (65%) |

### 3.2. Automated ICF coding

#### 3.2.1. Development experiments: identifying the best word embeddings

Figure 4 illustrates the results of development set experiments to identify the best word embedding features to use for coding Mobility and Self-Care/Domestic Life mentions. We evaluated MIMIC, NIHCC, SSA, and clinicalBERT embedding features for both classification and candidate selection approaches, with and without the Action oracle.

**Figure 4.**
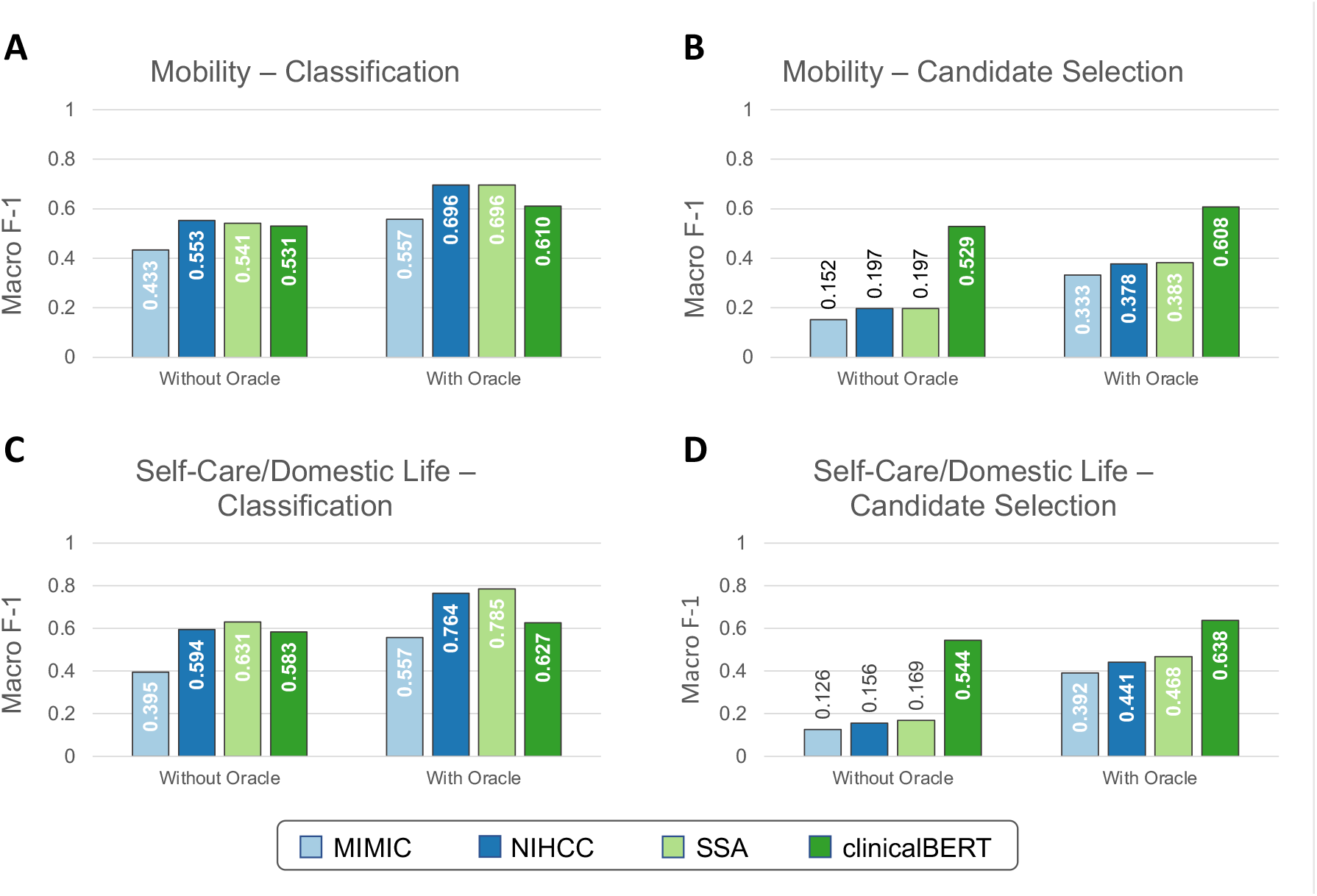
Development experiment results for selecting word embeddings. Development set performance (macro-averaged F-1 with ten-fold cross validation) is shown using each embedding strategy for both Mobility and Self-Care/Domestic Life data, using both classification and candidate selection approaches.

For the Mobility dataset, embeddings trained on the NIHCC and SSA corpora achieved highest development set performance both with the Action oracle (F-1=0.696 for both NIHCC and SSA) and without (NIHCC=0.553, SSA=0.541, difference not significant at *p*-value=0.9, bootstrap resampling). NIHCC embeddings were statistically significantly better than the next best clinicalBERT features (F-1 of 0.553 vs 0.531; *p-*value=0.025) without the Action oracle, while SSA embeddings were not significantly different from clinicalBERT (F-1 of 0.541 vs 0.531; *p*-value=0.17). We therefore took NIHCC embeddings as the best-performing features for classification experiments on the Mobility test set.

For the Self-Care/Domestic Life dataset, SSA embeddings achieved highest development set performance both with the Action oracle (SSA F-1=0.785 vs NIHCC F-1=0.764; *p*-value=0.031) and without (SSA=0.631, NIHCC=0.594; *p*-value=0.015). We therefore took SSA embeddings as the best-performing features for Self-Care/Domestic Life classification experiments.

Under the candidate selection approach, clinicalBERT features significantly (*p*≪0.001) outperformed all other embeddings on both datasets. We used clinicalBERT embeddings as the best-performing features for test set candidate selection experiments.

#### 3.2.2. Main experiments

Figure 5 shows overall performance of classification and candidate selection experiments on the Mobility and Self-Care/Domestic Life test sets. Classification models consistently outperformed candidate selection (*p*=0.041 for Mobility without Action oracle; *p*≪0.001 for Mobility with Action oracle and both settings of Self-Care/Domestic Life). This is consistent with our prior findings of comparable or slightly lower performance for our candidate selection model on Mobility data from physical therapy encounters (16). The Action oracle significantly (*p*≪0.001) improved performance in all cases, clearly demonstrating the value of building NLP systems to extract the Action components of activity mentions.

**Figure 5.**
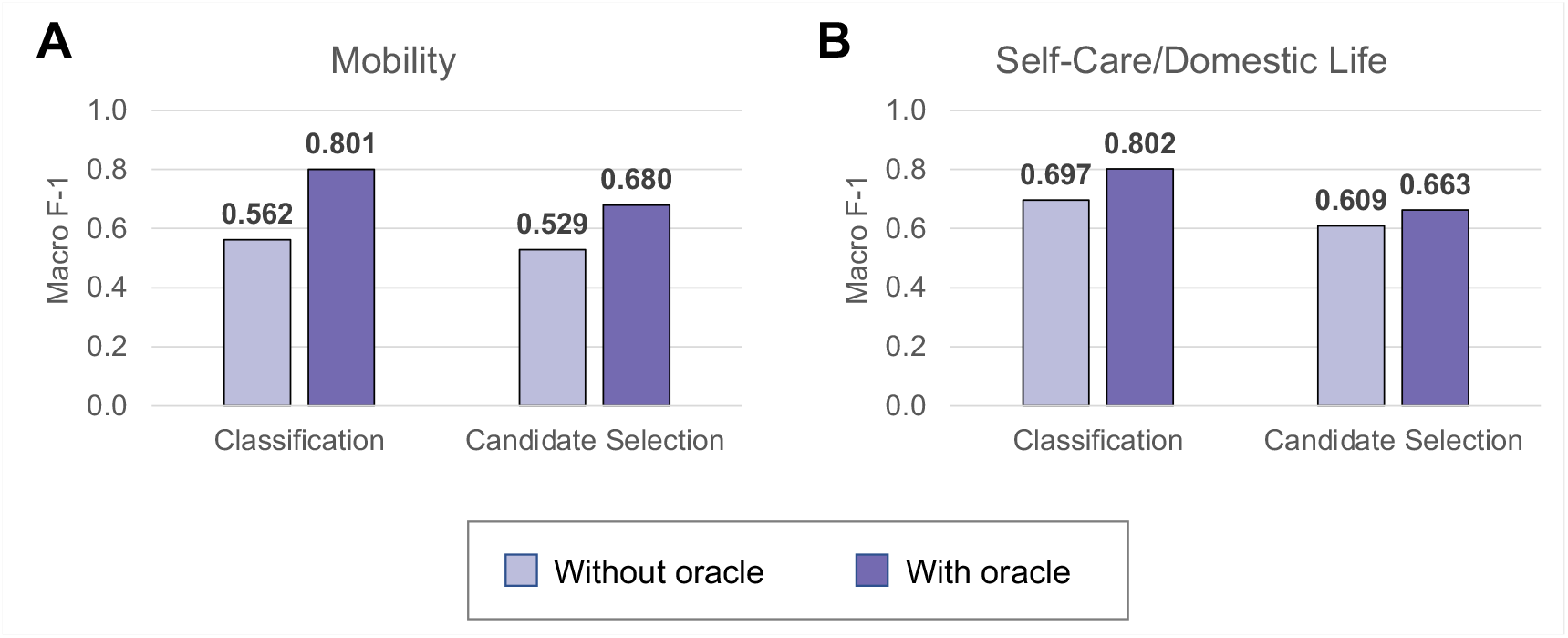
Test set performance on automated ICF coding in Mobility (Panel A) and Self-Care/Domestic Life (Panel B) test sets. Performance is reported for the best classification (Mobility: NIHCC embeddings; Self-Care/Domestic Life: SSA embeddings) and candidate selection (both datasets: clinicalBERT embeddings) models.

We further analyzed performance on each individual label in the Mobility dataset (shown in Figure 6) and the Self-Care/Domestic Life dataset (shown in Figure 7). Performance generally trended with the frequency of the label—i.e., both classification and candidate selection performance were best for the most frequent categories and gradually degrades for less frequent categories. We did not observe any categories where our classification or candidate selection models showed a clear advantage; rather, our classification models tended slightly higher than candidate selection on almost all categories. Having access to the location of Action components (i.e., with the Action oracle) improved performance on almost all categories, with most of the largest gains on rare categories: e.g., an F-1 gain of 0.25 (candidate selection) and 0.5 (classification) on d460 (21 samples) in Mobility data, and an F-1 gain of 0.3 (candidate selection) and 0.33 (classification) on d560 (22 samples) in Self-Care/Domestic Life data.

**Figure 6.**
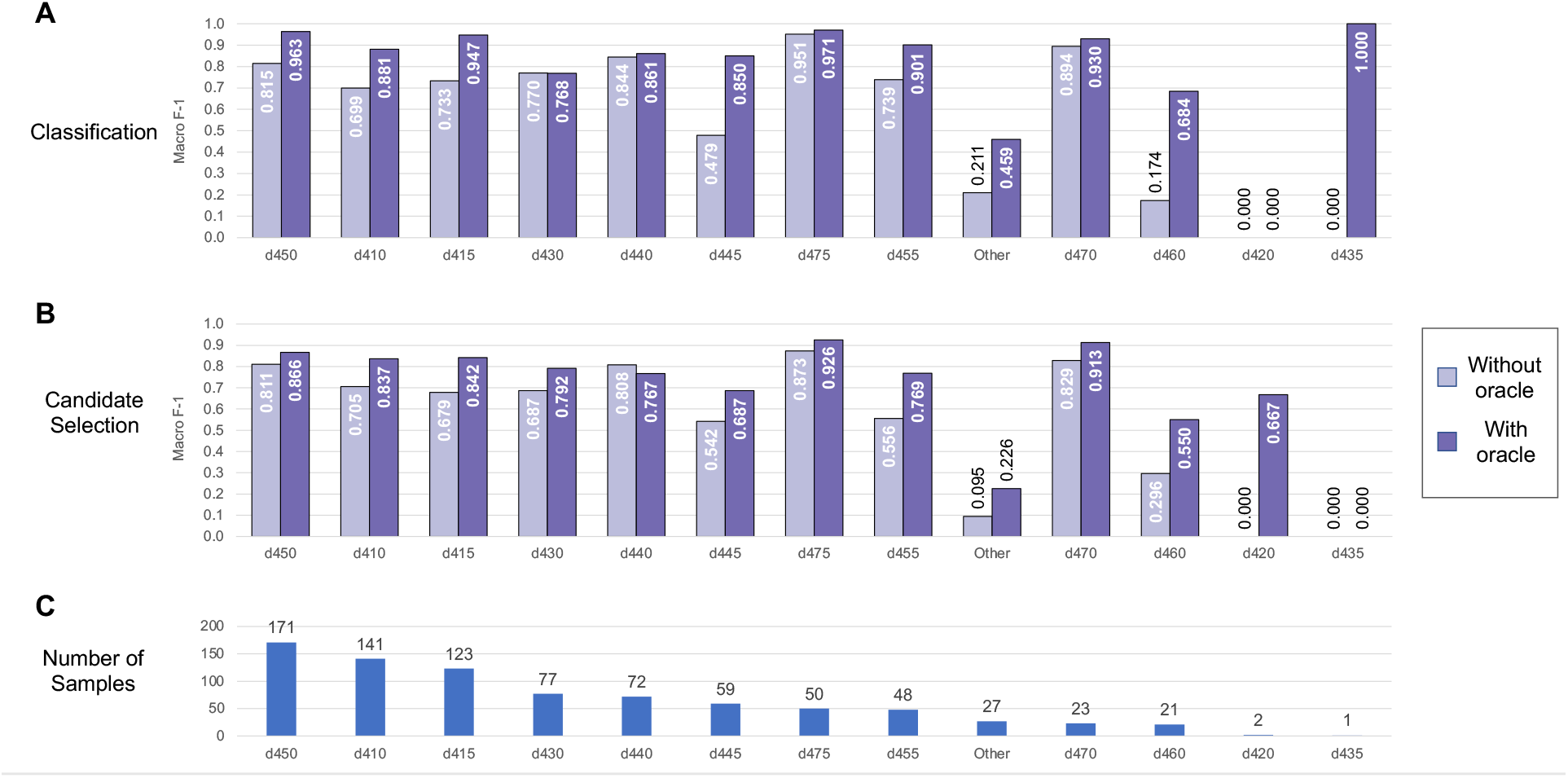
Automated coding performance for each distinct code in the Mobility dataset. Classification results are shown in Panel A, and candidate selection results in Panel B. Codes are ordered by descending frequency (illustrated in Panel C).

**Figure 7.**
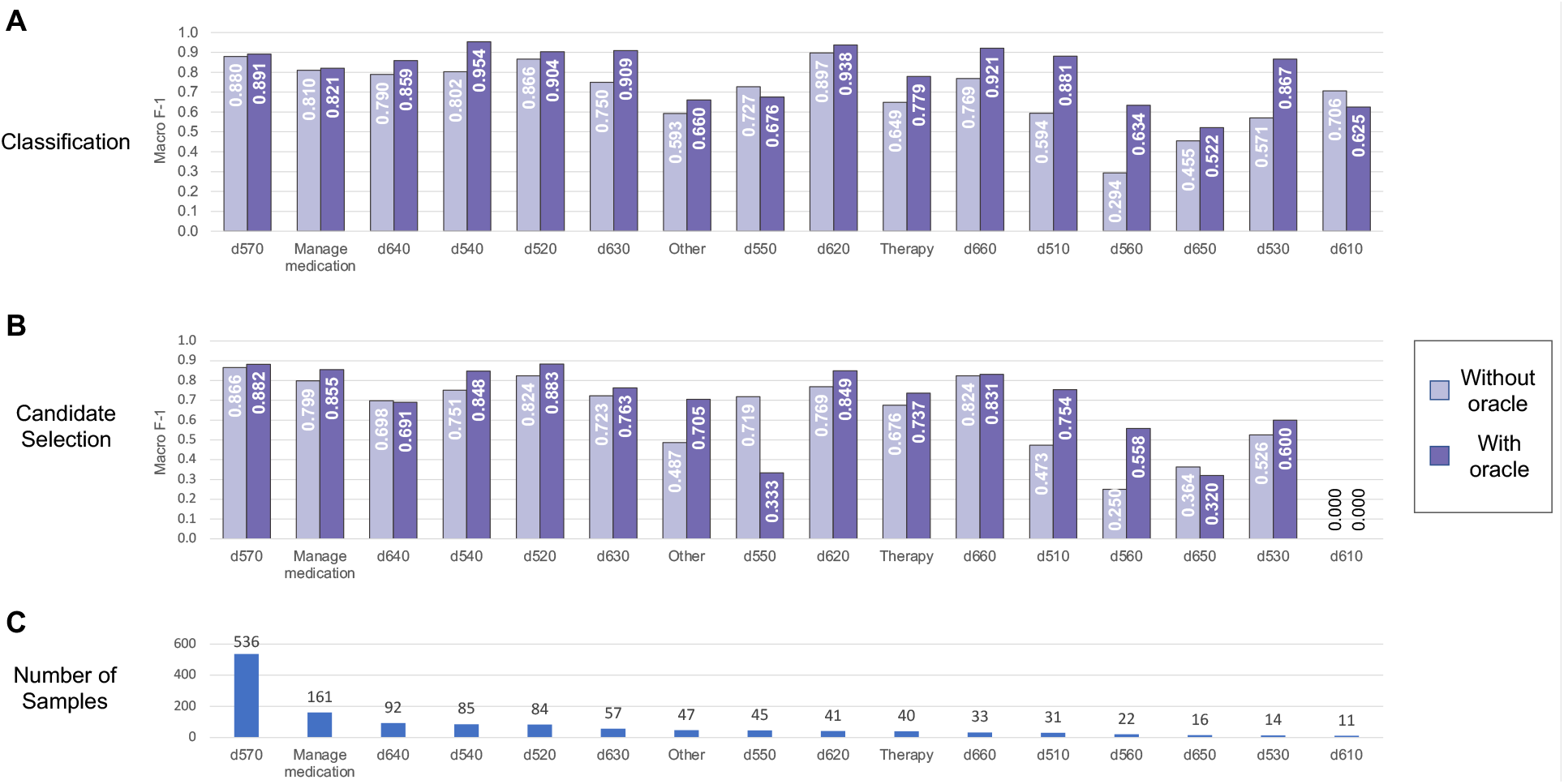
Automated coding performance for each distinct code in the Self-Care/Domestic Life dataset. Classification results are shown in Panel A, and candidate selection results in Panel B. Codes are ordered by descending frequency (illustrated in Panel C).

## 4. Discussion

We have shown that rich and diverse information on Mobility, Self-Care, and Domestic Life is recorded in free text health records collected from health providers by SSA for disability benefits adjudication. We presented NLP systems to map this information to specific ICF categories using two paradigms: classification (comparing each sample to other, previously-seen samples) and candidate selection (comparing a sample to ICF categories directly). Our experiments demonstrated that these systems show promising performance for enabling automated analysis of medical evidence through the lens of the ICF.

Our study also revealed limitations of the ICF as a practical tool for analyzing medical documentation. We discuss key insights from our annotation process in the following section, and highlight the particularly complex case of ICF category d570 *Looking after one’s health*. We further identify particular successes and challenges arising from our NLP experiments, and discuss implications of NLP tools for functional status—aligned to the ICF or to another conceptual framework—in both the SSA use case of disability adjudication and broader applications in clinical care and research.

### 4.1. Practical limitations of the ICF for Mobility, Self-Care, and Domestic Life information

Coding functional status information according to a standardized framework such as the ICF allows us to identify what kinds of functioning are discussed in health records and to organize information on patient functioning for retrieval and analysis. The ICF, as the internationally accepted classification of human functioning, is an important touchstone for this work, and it allowed us to capture a broad set of information about functional activity in free text health records. However, some activity mentions we observed in practice did not align with the categories presented in the ICF, such as “managing stairs”, “doing household tasks”, and “cleaning”. At the same time, other categories had significant overlap with one another in the expert annotation process, such as d450 *Walking*, d455 *Moving around*, and d460 *Moving around in different locations*. Category d465 *Moving around using equipment* was excluded entirely from annotation, as our information model represented assistive equipment (Assistance component) separately from the action being performed (Action component); this category therefore reduced to d455 *Moving around*. Some activity descriptors were highly context-dependent for selecting the appropriate ICF category: for example, we annotated “drinking” as d560 *Drinking* for the generic action of drinking, but as d570 *Looking after one’s health* when used to refer specifically to drinking alcohol (e.g., “He drinks two shots of whiskey a day”). Thus, while the ICF is clear and comprehensive for coding many Mobility, Self-Care, and Domestic Life activities, its use is often more theoretical than practical when applied to actual clinical reporting.

#### 4.1.1. ICF category d570 is overly broad

The limitations of the ICF in practice were particularly clear for the Self-Care category d570 *Looking after one’s health*. We found this category to be significantly over-represented in our data (accounting for 43.6% of all observed Self-Care/Domestic Life actions), and extremely broad in practice. Category d570 was treated as referring to preventative measures (e.g., exercising, taking prescribed medications, etc.) a person does to, or for, themselves or will/plans to do in the future. We excluded from consideration interventions performed or planned by healthcare providers, the goals providers set for themselves, and descriptions of specific therapy sessions that are not directly related to Self-Care. With this operational definition, we coded d570 for information as diverse as:

- She exercises 4 to 5 times a week
- Stretching, breathing techniques
- He drinks two shots of whiskey a day
- She has had two suicide attempts in the past
- He smokes a pack of cigarettes a day
- Takes over the counter supplements
- He is compliant with treatment but remains symptomatic
- I haven’t gone to counseling but I talk to my friend who is a preacher
- He consumed caffeine 1-2 times a week

Notably, we found category d570 in practice to include several social determinants of health, such as drug and alcohol use (also including misuse and abuse) and smoking status. In addition to the breadth of information, several activity mentions we coded with d570 required some level of inference on the part of the reader to understand the functioning described. For example, we annotated “I talk to my friend who is a preacher” in the example above as d570 because in the context of referring to counseling, this can be understood as the patient establishing a connection and/or reaching for help to look after themselves. References to suicide attempts were also coded as d570 because of the detriment involved to the physical and mental health of the patient.

From a practical standpoint in the annotation process, activity mentions coded with d570 presented two further complications. While stated (or implied) reasons for a patient taking care of themselves or not were not generally included in annotating activity mentions, in some cases they provided context to clarify whether an action was related to taking care of oneself or not. For example, in “her tendency to take a double shift knowing that there will be a detrimental impact on her comfort and health status”, the phrase “take a double shift” alone is not sufficient to determine a category of d570; including its effect on the patient’s health provides the necessary context to clarify that this is related to taking care of oneself. In addition, d570 was the only category where negation needed to be captured as part of the Action component, when it pertained to suicide or other self-harm, recreational drug and/or alcohol use, or medication non-compliance.

In summary, we found that the ICF is not necessarily in line with the types of information providers record about Self-Care, and that category d570 was too broad to effectively represent the diversity of Self-Care activities described in the data.

#### 4.1.2. Distinguishing patient engagement in “Therapy” and “Manage medications” from other uses of d570

We took the step in this study of specifically distinguishing patient engagement in *Therapy* (non-pharmacological) and *Manage medication* as distinct Self-Care categories, separate from the broader d570 category. We found that clinical notes frequently provided detailed information on how patients were or were not engaging actively in specific therapeutic interventions and determined that separate categories would provide a more organized view of the patient’s self-care activities as a whole. We distinguished between adherence to regimens for managing medications, which are therapies that a licensed provider needs to approve (in contrast to over-the-counter products such as multivitamins or alternative medicines, which we classified as d570), and participation in non-pharmacological therapies such as addiction treatment programs, physical therapy, occupational therapy, cognitive behavior modification therapy, psychological therapy and/or counseling, and anger management. To provide concrete examples of these distinctions, and further illustrate the complex scope of category d570, Table 5 (drawn from our annotation guideline (32)) presents a selection of samples for each label together with notes on why the information was or was not annotated as presented.

**Table 5.** Examples for the related labels of ICF category d570, Manage Medication, and Therapy. Brief notes are provided for each example as to why it was or was not annotated as shown. Activity mentions are indicated using yellow highlights, and Actions are indicated using underlines.

| Category | Examples | Notes |
| --- | --- | --- |
| d570 | Her sleep varies and she never feels rested | Not annotated; these fall within the Body Functions domain of the ICF. |
|  | <u>She has had a previous suicide attempt</u> | Suicidal actions are annotated as indicating risks to health. |
|  | <u>He drinks a six-pack of beer a day</u> | Reference to alcohol consumption. |
|  | <u>Patient was well-nourished</u> | Indicates the person is taking care of themselves. |
|  | <u>Her tendency to take a double shift knowing that there will be a detrimental effect on her comfort and health status</u> | Significant context needed to clarify the impact on self-care. |
| Manage Medication | He is currently prescribed medication by his neurologist to slow down the progression of his symptoms | Not annotated; does not state whether the person is actually taking the medications or not. |
|  | <u>Pt is currently on medication</u>: Prazosin at bedtime... | Medications the patient is currently taking; the medications themselves are not annotated. |
|  | <u>She takes Tylenol</u> | Reason for medication not needed; the specific medication is annotated to clarify what action is being performed. |
| Therapy | He has had no psychiatric care and no history of psychiatric hospitalization | Not annotated; reference to therapeutic care the patient has not used. |
|  | <u>She had occupational therapy</u> for a custom splint | Therapy for a particular purpose related to health. |
|  | <u>He was seeing a counselor</u> for his drug addiction | Counseling for a particular purpose related to health. |

#### 4.1.3. Overlap between d570 and other domains of the ICF

The interactions between health condition, body functions and structures, activities and participation, and contextual factors are at the heart of the ICF’s biopsychosocial model of human function. However, we found that particularly for category d570, both its definition and our observations of it in practice overlapped significantly with other domains of the ICF, creating an additional challenge for aligning clinical observations to the ICF model. Terms used in the definition of d570, such as “ensuring”, “appropriate level”, “avoiding harm”, and “being aware of the need” are more aligned with the b1 *Mental functions* heading in the Body Functions domain. At the same time, several examples we annotated as d570 included elements more in the domain of Personal Factors—these included references to work preferences, physical activity levels, etc. As the ICF does not currently classify Personal Factors, these elements cannot be classified separately from the activity of d570. However, alternative models can also inform approaches to representing these relationships in practice: for example, the Institute of Medicine’s 1997 model (33) separates personal factors into biologic factors (less modifiable) and lifestyle and behavior factors (more modifiable), and represents them as transitional factors in the enabling-disabling pathway. This perspective provides a framework for viewing the activity of *Looking after one’s health* as an outwardly observable act affected by internal processes such as personal health behaviors and choices. Modeling these relationships thus represents an important area of further inquiry both in refining the ICF model and in developing information technologies to align clinical observations with it.

#### 4.1.4. Implications for updating the ICF

Our findings suggest specific ways in which the ICF could be updated to decrease overlap between codes and better align with practical clinical reporting needs. Specific recommendations supported by our analysis include: (1) Remove the term “walking” from the definition of d460 *Moving around in different locations*, to reduce overlap with d450 *Walking*. (2) Explicitly distinguish between the general action of drinking liquids, represented by category d560 *Drinking*, and the specific case of drinking alcohol (which providers often refer to simply using “drinking” or “drinks”, e.g., “his drinking habit” or “two drinks nightly”), which overlaps with d570 *Looking after one’s health*. (3) Replace the broad category d570 *Looking after one’s health* with multiple, more specific categories that reflect particular behavioral patterns, such as physical or cognitive exercises, substance use (ordered or disordered), or treatment compliance.

### 4.2. NLP is a promising technology for analyzing FSI in clinical free text

Our experiments demonstrate that NLP technologies can help to organize FSI in free text portions of the medical record, making this information easier to find and use in decision making processes. Our findings identify particular opportunities for future work on refining and expanding these technologies, and we further discuss potential implications of these technologies in managing SSA disability programs as well as individual patient care.

#### 4.2.1. Successes and challenges in automated ICF coding with NLP

The NLP systems developed in this work achieved high performance for the majority of Mobility and Self-Care/Domestic Life ICF categories. The Action oracle was the single largest factor in system performance—F-1 on Mobility codes increased by 0.22 on average for classification and 0.15 on average for candidate selection; increases for Self-Care/Domestic Life were smaller but still considerable at 0.11 average for classification and 0.05 average for candidate selection. The first step in further refining NLP methods for analyzing FSI must therefore be to include identification of Action components in the process of extracting activity mentions from text.On a per-category basis, the best NLP models achieved high performance for most ICF categories. In Mobility, we achieved over 0.9 F-1 for five high-impact categories: d450 *Walking*, d415 *Maintaining a body position*, d475 *Driving*, d455 *Moving around*, d470 *Using transportation*. (d435 *Moving objects with lower extremities* is not included in this list as only one sample was present in the test set, limiting the reliability of performance evaluations for this category). In Self-Care/Domestic Life, we exceeded 0.9 F-1 for five common categories: d540 *Dressing*, d520 *Caring for body parts*, d630 *Preparing meals*, d620 *Acquisition of goods and services*, and d660 *Assisting others*. System performance was not strongly correlated with the frequency of the ICF categories, indicating that in most cases, there is a clear separation between categories. However, many of the errors made by all systems were mispredictions of the most frequent labels (d450 *Walking* for Mobility, d570 *Looking after one’s health* for Self-Care/Domestic Life); frequency effects are thus still an important issue to address in further refinement of NLP models for ICF coding.

Per-category performance was more consistent for Self-Care/Domestic Life than for Mobility, despite the higher skew of the Self-Care/Domestic Life category distribution; this may reflect greater issues of category overlap in the Mobility domain. In both Mobility and Self-Care/Domestic Life data, the *Other* category was a consistent challenge, reflecting its nature as a catch-all category for samples that could not be mapped cleanly to single categories in the ICF.

#### 4.2.2. Potential applications in the SSA disability adjudication process

The process of adjudicating applications to the SSA for federal disability benefits was one of the motivating use cases for this study. The adjudication process includes collection and review of highly heterogeneous medical evidence, frequently collected as free text or semi-structured documents, to identify whether a person meets the necessary criteria for determining disability. This is a sequential process, which involves identifying information related to functioning at multiple steps. Claimants may be allowed based on meeting specified medical criteria organized into different body systems (34), where musculoskeletal criteria refer to several aspects of Mobility, criteria for mental disorders involve multiple areas of daily functioning, and criteria for multiple body systems refer to adherence to treatment. Claimants will also often report on daily activities and routines to provide details on functional abilities and limitations relevant to the workplace. Functional assessment is also a regular part of the adjudication process to determine whether a claimant is able to work, including through Residual Functional Capacity assessments which include physical assessments highly dependent on Mobility. Thus, NLP-based tools to extract information related to functioning and organize it according to a standardized framework such as the ICF could be of use at multiple points in the disability adjudication process (35).

#### 4.2.3. Broader implications of ICF coding with NLP

NLP systems like the ones developed in this study have significant potential for helping to advance both clinical research and patient care. Identifying and organizing the rich information on individual function currently locked away in medical free text can unlock valuable details to enrich researchers’ understanding of rehabilitation outcomes, and highlight salient details of patients’ experiences in clinical decision making. Prior research on automated and semi-automated ICD coding systems using NLP methods provides an instructive example of how these approaches can streamline medical coding processes (36–38). Growing integration of the ICF into clinical and research settings, from primary care (39) and EHR implementation (40) to pediatric research (41), present similar opportunities to smooth the adoption and practical use of ICF categories with NLP-based coding systems. Vreeman and Richoz (42) describe potential benefits to both clinical care and research from integrating the ICF and other standardized vocabularies into EHRs, and Bettger et al. (43) highlightthe role of EHR data in providing key insights to advance quality measures, research, and policy for rehabilitation. NLP technologies for ICF coding can serve as a valuable method to leverage the ICF as a lens to study the rich information collected in EHR notes.

In patient care, further development of NLP technologies can facilitate the decision-making process in several ways. Manabe et al. (44) developed an interactive system for selecting ICF categories in the EHR for mental health care; combining such an approach with NLP-based analysis could enable context-sensitive ICF coding during clinical note entry, improving the depth of information entered and its alignment with the ICF. At a new patient visit, NLP analysis of previously entered notes could also be used to highlight past limitations the patient experienced and inform patient-provider communication. Beyond the clinical setting, use of NLP technologies for social support programs (such as the SSA disability programs that motivated our study) can help to more rapidly identify and organize key information from an individual’s history to inform benefits decisions. Developing and evaluating new NLP technologies targeting further use cases in clinical research and patient care is a key direction for future research with significant potential for impact.

### 4.3. Limitations

The SSA documents used in this study were a mix of clinical records sourced from healthcare providers around the U.S. and specialty records for consultations commissioned by SSA pertaining to a disability benefits claim. These documents are thus not representative of EHR notes in most health systems. In addition, the population who is the subject of these documents consists of claimants for federal disability benefits due to work-related disability; this population is not necessarily representative of persons receiving rehabilitation care (or other care involving functional assessment) more broadly. From a practical standpoint, many of the SSA documents used exhibited severe noise from the OCR conversion process from scanned images to text. In our experiments, model design hyperparameters were not explored, nor were alternative classification or candidate selection methods, potentially limiting the F-1 measures we were able to achieve.

## 5. Conclusions

Valuable information about patient functioning is regularly recorded in the free text portions of the EHR. The expressivity of natural language allows for documentation of rich details about functional experience, from levels of functional limitations experienced in different contexts to the patient’s goals and priorities for their own functioning. While free text documentation is difficult to analyze with traditional methods, NLP technologies enable powerful, semantically-enriched analysis of functioning information without losing expressivity. We analyzed two datasets of clinical records pertaining to disability benefits claims submitted to the U.S. Social Security Administration, using the ICF to identify and organize documented information about claimants’ Mobility, Self-Care, and Domestic Life functioning. We found a rich diversity of functional status information in SSA documents, and developed NLP models to automatically code this information according to the ICF. Our models achieved strong performance across key types of Mobility, Self-Care, and Domestic Life activities, demonstrating promise for automatically organizing functional status information within the ICF framework for easier analysis and review. We identified several practical limitations of the ICF for coding clinical reports, particularly the overly broad formulation of the Self-Care category d570 *Looking after one’s health*. The results of this study and the NLP technologies assessed have significant implications for deepening the analysis of free text EHR data through an ICF lens, and will contribute to ongoing efforts to learn more from the EHR in rehabilitation.

## Data Availability

The datasets presented in this article include identified medical information collected by the U.S. Social Security Administration for the purposes of adjudicating claims for disability benefits, and are not able to be shared.

## Author Contributions

DNG: conceptualization of study, development of methodology, conducting experiments, data analysis, and lead author of this manuscript. JCM: development of methodology, data collection and annotation, co-author of this manuscript. PSH: development of methodology, data collection and annotation, co-author of this manuscript. MS: development of methodology, data collection and annotation. RJS: development of methodology, data collection and annotation, statistical analysis. JP: project administration, development of methodology, data collection, co-author of this manuscript. LC: acquisition of funding, project administration. All authors contributed to this article and approved the submitted version.

## List of Abbreviations

EHR: electronic health record
FSI: functional status information
IAA: inter-annotator agreement
ICF: International Classification of Functioning, Disability and Health
NLP: natural language processing
SSA: U.S. Social Security Administration

## Acknowledgments

We thank Chunxiao Zhou and Alex Marr for invaluable assistance in data management and calculation of inter-annotator agreement. We also gratefully thank Elizabeth Rasch for invaluable discussions and feedback on this article.

## Ethics Statement

Ethics review was performed by the NIH Clinical Center Office of Human Subjects Research, and this research was determined to meet the criteria for not human subjects research.

